# Analysis of fatality among COVID-19 cases in Mexican pregnant women: a cross-sectional study

**DOI:** 10.1101/2022.10.12.22280996

**Authors:** Nicolás Padilla-Raygoza, María de Jesús Gallardo-Luna, Gilberto Flores-Vargas, Efrain Navarro-Olivos, Francisco J. Magos-Vázquez, Elia Lara-Lona, Daniel A. Díaz-Martínez

**Author notes:** Corresponding Author (NP-R). Department of Research and Technological Development, Víctor Cervera Pacheco #14 Primer Piso, Plaza Santa Fé, Guanajuato, Gto., Mexico (Nicolás Padilla-Raygoza). Directorate of Teaching and Research, Víctor Cervera Pacheco #14 Primer Piso, Plaza Santa Fé, Guanajuato, Gto., Mexico CP36250 (Efrain Navarro-Olivos). Directorate of Health Services, Tamazuca #4, Guanajuato, Gto., Mexico CP36000 (Francisco J. Magos-Vázquez, Elia Lara-Lona, Daniel A. Díaz-Martínez).

## Abstract

This study aims to analyze the fatality of cases confirmed by COVID-19 among pregnant women in Mexico. It is a cross-sectional and analytical study. We used the registries from pregnant women available in the open database of the National Epidemiological Surveillance System from the General Directorate of Epidemiology. We showed descriptive statistics for all the variables. A suspected case of COVID-19 is any person who presented the following signs and symptoms: fever, headache, cough, and others. A confirmed case is any suspected case with a positive RT-PCR test result. We computed OR and 95% confidence intervals to estimate the effect of independent variables on dying from COVID-19. Also, it was calculated the Case Fatality Ratio (CFR) among pregnant women. The alpha value was fixed at 0.05 as a threshold to show statistical significance. The CFR was 1.09%. For confirmed cases, the average age among those who died was higher than among those who did not die (P <0.05). The average time between the onset of symptoms and registration in the system was higher for those who died (P <0.05). Among the deceased, 76.97% had pneumonia. For the 40-49 years age group, the effect on death was statistically significant (OR 4.97, CI95% 1.77 – 17.85). Outpatient care had a protective effect on dying (OR 0.04, CI95% 0.02 – 0.09). Pneumonia was highly associated with death (OR 8.68, CI95% 5.72 – 13.6). Co-morbidities did not affect dying while considering them in a multivariable logistic regression model. Among pregnant women, smoking has little effect on death by COVID-19. The CFR was low compared with the rest of the Mexican population. The co-morbidities had a low prevalence among pregnant women. Since the reproductive age span is young age, pregnant women have two protective factors for COVID-19 detected so far: being young and woman.

## Introduction

In November-December 2019, several cases of pneumonia of unknown origin were detected in Wuhan, China [1]. After genomic analyses, the pathogen responsible was determined and called Severe Acute Respiratory Syndrome Coronavirus 2 (SARS-CoV-2). Meanwhile, the disease was named Coronavirus Disease 2019 (COVID-19) [2]. Since then, this disease has spread to the entire world. By November 1, 2021, 246,594,191 cases and 4,998,784 deaths have been reported in 224 countries [3].

Pregnant women are likely to represent a high-risk population in the COVID-19 pandemic. Although pregnant women infected with SARS-CoV-2 are predominantly asymptomatic or present mild COVID-19 -very similar to non-pregnant women [4]-they may be at higher risk of severe COVID-19 than the general population, according to Wastnedge et al. [5].

In a report from seven pregnant women infected by SARS-CoV-2 in Wuhan, China, all were in the last pregnancy trimester and had contact with the epidemic area, but not with food from the seafood market in the same city. The median age was 32 years -spreading between 29 and 34 years- and none was required to be treated in the ICU [6].

Pregnant women are considered a group at risk during the pandemic by SARS-CoV-2 due to biological processes related to pregnancy. It is hypothesized that a shift in the CD4 + cell population towards the Th2 phenotype, promoting the humor response, mediates the altered inflammatory response to viruses during pregnancy. The Th2 and Th1 response has been implicated in the pathogenesis of severe COVID-19 [7]. Also, during pregnancy, a decrease in natural killer (NK) cells occurs [8], and the NKs participate in virus neutralization [9]. The effect of the NK decreasing in the clinical picture of COVID-19 is unknown. There are other immune effects as progesterone levels [10] and alterations in Toll-like receptors (TLRs) [11]. It remains to be elucidated whether they have protective or harmful effects on pregnant women with COVID-19 [5]. Besides, the reduction in total lung capacity and the inability to expel secretions can make pregnant women more susceptible to severe respiratory infections [12].

Pregnancy is also a hypercoagulable state, with increased thrombin production and intravascular inflammation [13]. On the other hand, COVID-19 has been associated with thromboembolic complications in 31% of the patients reported in Knight M et al. [14].

According to Lopez-Rodriguez et al. [15], in 29,416 pregnant Mexican women, 39% had a COVID-19-positive diagnosis. They found that the risk of death was three times higher among those positive for COVID-19 than those who tested negative.

This paper aims to analyze the cases of COVID-19 in pregnant women registered in the open and public database of the National Epidemiological Surveillance System (NESS) from the General Directorate of Epidemiology (GDE) of the Ministry of Health (MH) [16] and their association with comorbidities.

## Material and methods

The study was approved by the Research Ethics Committee from the Salamanca General Hospital, Mexico.

### 2.1 Study design

It is a cross-sectional and analytical study.

As a requisite to be tested, pregnant women must meet the operational definition of suspected cases of respiratory viral infection (17). A suspected case of viral disease is considered as anyone who presents fever, headache, cough, or dyspnea (signs of severity), in addition to one of the following: myalgias, arthralgias, odynophagia, chills, chest pain, rhinorrhea, anosmia, dysgeusia, or conjunctivitis. If a subject meets the operational definition, the subject undergoes a nasopharyngeal Real-Time Polymerase Chain Reaction (RT-PCR) test (17).

The variables included in the database are the following: state of residence, age, date of onset of symptoms, date of registration, date of death (if it occurred), hospital or outpatient management, pneumonia diagnosis, underlying presence of diabetes, Chronic Obstructive Pulmonary Disease (COPD), asthma, immunosuppression, hypertension, cardiovascular disease, chronic kidney disease, obesity, or smoking. The diagnosis tool for COVID-19 was the real-time polymerase chain reaction test (RT-PCR) (16, 17).

The database does not record gestational age at the time of positive diagnosis for SARS-CoV-2.

After the approval of the research protocol by the ethics committee for research, we downloaded the open database of the NESS/GDE, and we preprocessed it to verify the data consistency. The dataset was analyzed in STATA 13.0 ® (Stata Corp., College Station, TX, USA). NESS/GDE database does not include personal data.

### 2.2 Statistical analysis

Descriptive statistics are presented for all the variables. To compare quantitative variables between confirmed COVID-19 cases, deceased or not, the Student t-test was performed. To compare categorical variables, the Chi-squared was used. Odds Ratio (OR) and 95% confidence intervals (95%CI) were calculated to estimate the effect of independent variables on dying from COVID-19. Also, it was calculated the Case Fatality Ratio (CFR) among pregnant women. In all cases, the corresponding P-values are present. Also, to demonstrate the statistical significance of the results, the alpha value was set at 0.05. Statistical analysis was performed in STATA 13.0 ® (Stata Corp., College Station, TX, USA).

## Results

### Description of the pregnant women population

In the NESS/GDE database, 42,334 pregnant women were reported; from them, 13,891 (32.81%) tested positive for SARS-CoV-2 and 28,443 (67.19%) tested negative (16).

### Characterization of death cases

Both SARS-CoV-2 positive and negative pregnant women had an age range of 12 to 49 years and a mean of 28.41 ± 6.13 years for the former and 27.09 ± 6.36 years for the latter -the difference was statistically significant (P = .00001)-. According to the database, the difference between the onset of symptoms and registry into the system, on average, is higher among COVID-19 cases than among negative ones (P <.05) (Table 1).

**Table 1.** Distribution of quantitative variables among deaths and non-deaths by COVID-19 in Mexican pregnant.

| Variable | Confirmed COVID-19 cases |  | t-test, df, *P*-value |
| --- | --- | --- | --- |
|  | Deaths<br>(n=152) | Non-deaths<br>(n=13,739) |  |
| Age (years) |  |  |  |
| Minimum to maximum | 16 to 49 | 12 to 49 | 6.42, 13889, <0.001 |
| Mean ± S | 31.59 ± 6.89 | 28.38 ± 6.12 | |
| Time between beginning of symptoms and date of registry (days) |  |  |  |
| Minimum to maximum | 0 to 16 | 0 to 39 | 4.96, 13899, <0.001 |
| Mean ± S | 4.90 ± 3.55 | 3.68 ± 3.06 | |

**Abbreviations:** df degree of freedom

The CFR was 152/13,739 (1.1%). For confirmed cases, the average age among the deceased cases was greater than among those who did not die (P <0.05). The average time between the onset of symptoms and registration into the system was greater among those who died. (P <0.05) (Table 1).

Between pregnant women positive for COVID-19, the CFR by age group were: 12-19 years, 0.44%, 20-29 years, 0.78%, 30-39 years, 1.36%, and 40-49 years, 3.74%.

The predominant age group among the deceased cases was 30-39 years (47.37%).

Meanwhile, in the non-deceased, those aged 20 to 29 years (51.63%) (P <0.05) predominated.

### Associated risk factors for lethality

Among the deceased cases, hospitalized patients predominated (93.42%). Compared with the non-deceased, where most subjects received outpatient management (82.27%) (P <0.05). Among the deceased, 76.97% had pneumonia. Among the non-deceased, only 17.73% had it (P <0.05). A statistically significant relationship with death (P <0.05) was found for diabetes, COPD, asthma, immunosuppression, hypertension, chronic kidney disease, obesity, and smoking (Table 2).

**Table 2.**
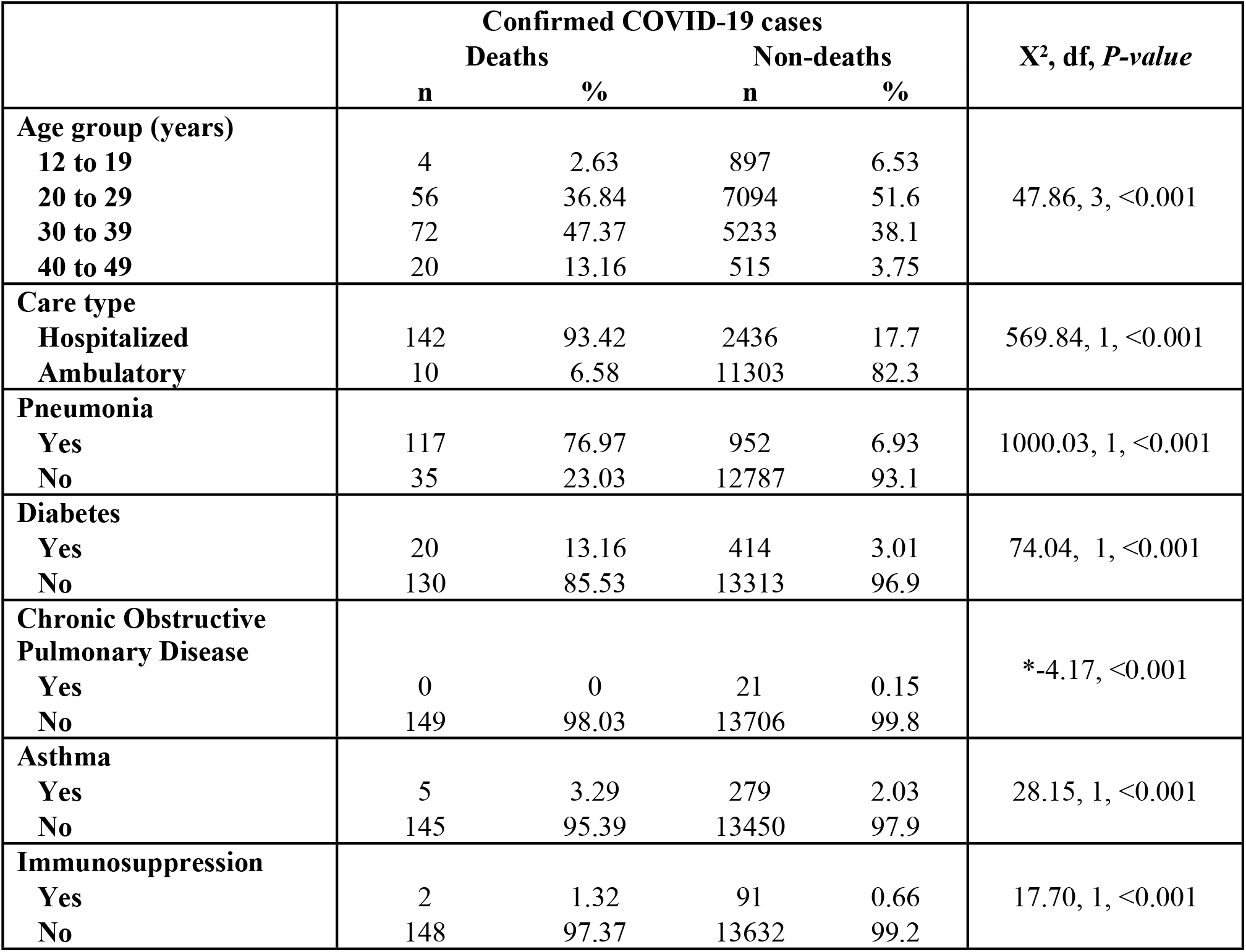
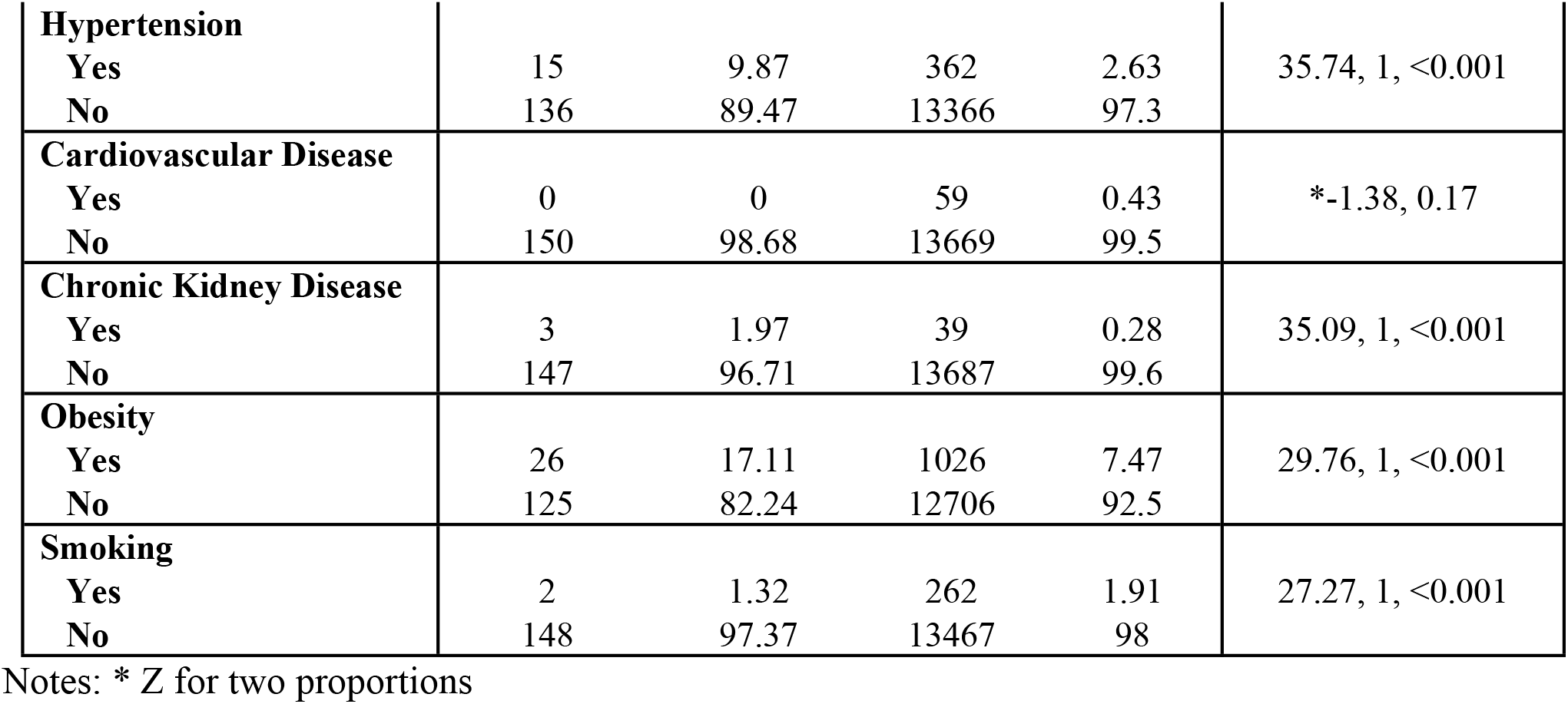
Relationship between variables and deaths by COVID-19 in Mexican pregnant women.

Using the logistic regression model that included all the variables with a statistically significant relationship with death, an effect on dying was detected for the age group from 40 to 49 years (OR 4.97 95%CI 1.77-17.85). The difference in days between the onset of symptoms and system registration is in the border area. The type of outpatient care has a protective effect on dying (OR 0.04 95%CI 0.02-0.09). Meanwhile, Pneumonia shows a high association (OR 8.68 95%CI 5.72-13.6). Diabetes, asthma, immunosuppression, hypertension, chronic kidney disease, and obesity did not affect death. As seen in the variance inflation factor (VIF), these variables may be highly correlated. Hence, they have little influence on the model. Smoking has little effect on death due to COVID-19 among pregnant women (Table 3). We fitted a model by omitting diabetes, immunosuppression, hypertension, and chronic kidney disease -those with higher VIF, different age groups, and non-statistically significant effects-. The Bayesian Information Criteria decreased from 1217.52 in the original model to 1190.26. The likelihood ratio test showed a statistically significant difference between the models. Nevertheless, the qualitative results were the same.

**Table 3.**
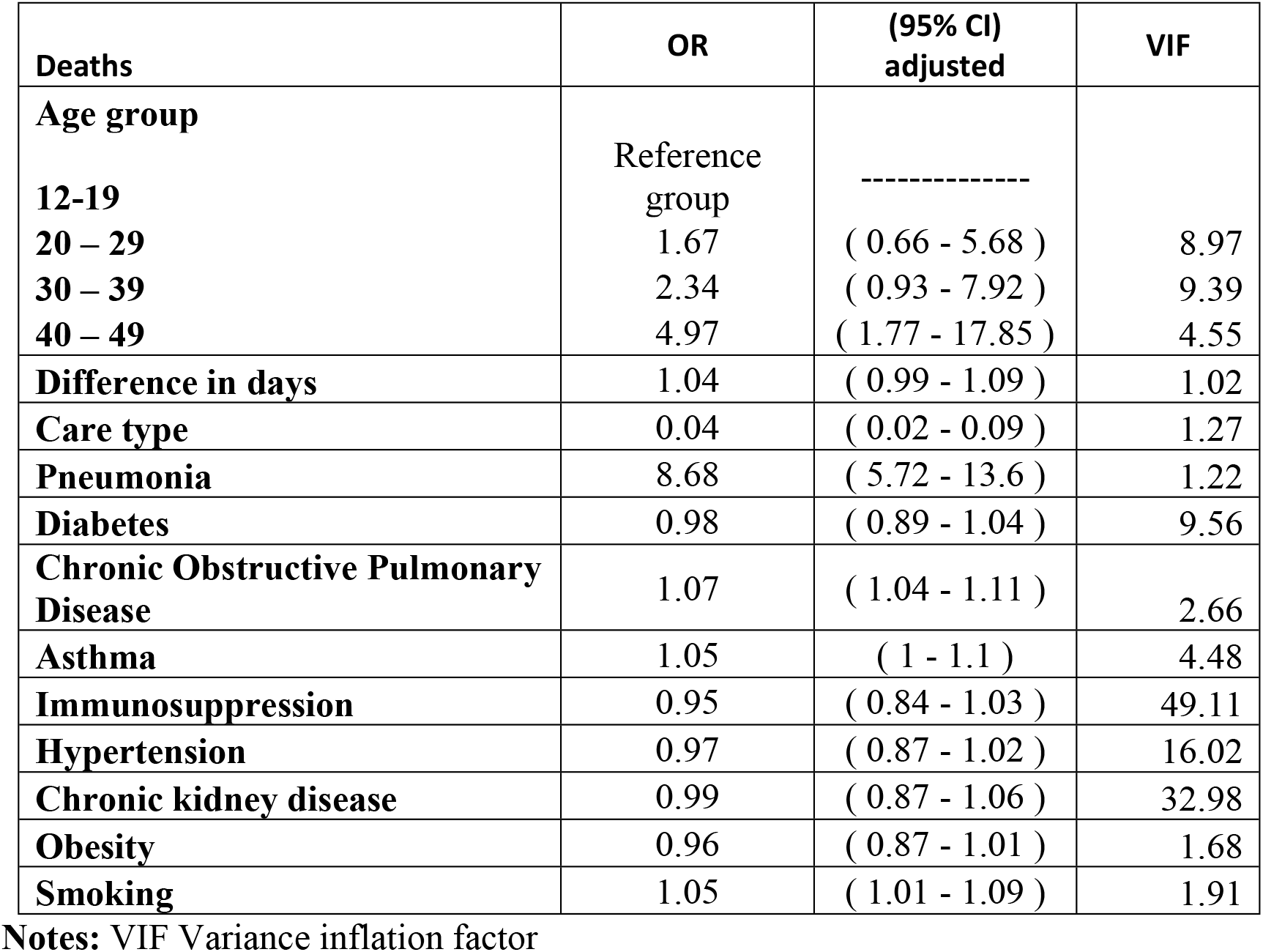
Logistic regression model including all independent variables with death in Mexican pregnant women.

## Discussion

The CFR reported by Padilla et al. [18] for the Mexican population on May 15, 2020 (10.59%) was higher than the one for the sample in this study (1.1%). Padilla et al. [18] also mentions that 68.14% of the deaths were in males and 49.57% in those over 60 years.

Table 2 reports a relationship between the study variables and death from COVID-19, except for cardiovascular disease. When quantifying the effect of these variables, only the age group, the difference in days between the onset of symptoms and the registration in the system, the type of care, pneumonia, COPD, asthma, and smoking affected death (Table 3).

Of nine pregnant women with COVID-19 and pneumonia, none developed severe pneumonia or died, as reported by Cheng et al. [19]. In the sample of Mexican pregnant women, 18.56% were hospitalized, 7.70% developed pneumonia, and 1.09% died (Table 2). In a series of 116 cases of pregnant women with COVID-19, 8% developed pneumonia [20], like the results in this study. Yu et al. [21] reported 124 maternal deaths in Brazil between February and June 2020. At the same time, Mexico reported seven maternal deaths [22]. The increase in maternal mortality from COVID-19 in Brazil may be due to a decreased availability of health professionals or a lack of intensive care units.

There is no evidence that pregnant women are more susceptible to SARS-CoV-2 [23].

In the analysis of the relationship between different variables with dying from COVID-19 was found that age group, hospitalization, pneumonia, diabetes, COPD, asthma, immunosuppression, hypertension, chronic kidney disease, obesity, and smoking are related to death from COVID-19 (P <0.05) (Table 2). Nevertheless, these relationships are lost when analyzing the logistic regression model that included all the variables associated with death from COVID-19.

The age group of 40-49 years shows an effect on dying from COVID-19 (OR = 4.97, 95% CI 1.61 -15.36). The age group from 40 to 49 years had the higher CFR, 3.74%.

The difference in days between the onset of symptoms and the date of registry in the NESS/DGE system is at the limit of statistical significance (OR = 1.04, 95% CI 1.00-1.09). Pneumonia is the one variable with the highest effect on dying (OR = 8.68, 95% CI 5.64 - 13.37). COPD, asthma, and smoking show ORs close to 1 (Table 3), which indicates a slight effect.

Pneumonia showed the highest OR on death from COVID-19 (Table 3), consistent with previous reports concerning children and adults, where the principal cause of death was pneumonia.

The underlying pathologies that aggravate COVID-19 in the general population seem to have minor effects on the severity of COVID-19 in pregnant women.

Pregnant women have been considered a risk group for COVID-19. In the database analyzed, the co-morbidities did not play a statistically significant role in death while integrating them in a multivariable logistic regression model, except for COPD and asthma. Also, health care workers must monitor the development of pneumonia because it is significantly related to death. A substantial limitation of this study is the absence of data about gestational age. This variable could play a confounding role in mortality. It is worth noting that some deaths occurred among pregnant women with ambulatory care. It should be addressed in further research. Due to the limitations of the database, it was not possible in this study.

## Conclusion

As in the general population, older age and pneumonia relate to death. The co-morbidities had a low prevalence among pregnant women. Since the reproductive age usually ends at 50, pregnant women have two protective factors for COVID-19 detected in the literature: being young and woman. They have been two inherent confounding factors in this study.

## Data Availability

The datasets analyzed for this study can be found in the Open Science Framework Padilla-Raygoza N. COVID-19 in Mexican pregnant [Internet]. OSF 2021. Available from: https://osf.io/b8hyw/

## Data Availability Statement

The datasets analyzed for this study can be found in the Open Science Framework Padilla-Raygoza N. COVID-19 in Mexican pregnant [Internet]. OSF; 2021. Available from: https://osf.io/b8hyw/

## Notes

### Competing Interest Statement

The authors have declared no competing interest.

### Funding Statement

Non funding for this research

### Author Declarations

The protocol was approved by Ethics Commite on Research of General Hospital of Salamanca (México)

